# SARS-CoV-2 antibody changes in patients receiving COVID-19 convalescent plasma from normal and vaccinated donors

**DOI:** 10.1101/2021.07.30.21261339

**Authors:** Judith Leon, Anna E. Merrill, Kai Rogers, Julie Kurt, Spencer Dempewolf, Alexandra Ehlers, J. Brooks Jackson, C. Michael Knudson

**Author notes:** Corresponding Author: C. Michael Knudson, Department of Pathology, University of Iowa Hospitals and Clinics, 200 Hawkins Dr., C250 GH.

## Abstract

**Background and Objectives:** Vaccination has been shown to stimulate remarkably high antibody levels in donors who have recovered from COVID-19. Our objective was to examine patient antibody responses following COVID-19 Convalescent Plasma (CCP) transfusion and compare responses to CCP from vaccinated and nonvaccinated donors.

**Materials and methods:** Plasma samples were obtained from 25 recipients of CCP and COVID-19 antibody levels measured before and after CCP treatment. Factors that effect antibody levels were examined.

**Results:** In the 21 patients who received CCP from nonvaccinated donors, only modest increases in antibody levels were observed. Patients who received two units were more likely to seroconvert than those receiving just one unit. The strongest predictor of changes in patient antibody level was the CCP dose. Using patient plasma volume and donor antibody level, the post-transfusion antibody level could be predicted with remarkable accuracy. In contrast, the 4 patients who received CCP from vaccinated donors all had dramatic increases in antibody levels following transfusion of a single unit. In this subset of recipients, antibody levels observed after transfusion of CCP were comparable to those seen in donors who had fully recovered from COVID-19.

**Conclusion:** If available, CCP from vaccinated donors with very high antibody levels should be used. CCP from vaccinated donors increases patient antibody levels much more than 1 or 2 units of CCP from unvaccinated donors.

## Introduction

The emergence of SARS-CoV-2, the causative agent of COVID-19, has resulted in intense efforts to identify new and effective treatments. The lack of clinically validated anti-viral therapies against coronaviruses led to the broad utilization of COVID-19 convalescent plasma (CCP) obtained from survivors of COVID-19 to treat patients with active disease [1-3]. While the mechanism of action of CCP is uncertain, the most prevalent hypothesis is that CCP contains neutralizing antibodies that limit viral spread and replication [4]. Multiple reports describe the rationale for this therapy and several studies provide some evidence of efficacy [5-10]. However, other trials have failed to show the benefit of CCP in hospitalized patients [11] and meta-analyses to date have drawn equivocal conclusions about the efficacy of CCP [12-14]. The closing of both the REMAP-CAP trial (NCT02735707) and the RECOVERY trial (NCT04381936), halted due to futility, have greatly dampened enthusiasm for CCP, however, it continues to be used for select patient populations. One common limitation in these trials is that patient SARS-CoV 2 antibody levels were not measured before enrollment, with retrospective antibody testing identifying many patients who were seropositive before treatment [11]. Antibody responses following CCP transfusion has not been broadly studied and when it has been studied the changes in antibody levels are often modest. While not a randomized trial, results from the expanded access protocol in the US (NCT04338360) demonstrate that hospitalized patients receiving high titer CCP had improved survival when compared to the low titer group [15]. Furthermore, recent reports have demonstrated that CCP donors who are vaccinated have dramatically higher spike-specific antibody levels with high neutralization titers than those that experience a natural infection [16, 17]. Given this information, one could hypothesize that patients receiving high quality CCP from vaccinated donors may be more likely to respond clinically.

In the early days of the current pandemic groups around the world rushed to utilize convalescent plasma obtained from COVID-19 survivors without clear guidelines for the selection of the best candidates for donation of CCP or identifying clinical parameters for patients most likely to benefit from CCP. This approach did not allow for early phase clinical trials that would look at the pharmacokinetics of the intervention in question and help with the dosing and timing of CCP as trials were designed. As a CCP donor program was established in our hospital-based donor center, nearly all the CCP used at our hospital came from donors with known antibody levels. To begin to identify what makes some CCP units more effective than others, an IRB-approved protocol to collect and store plasma on CCP recipients before and after they were transfused was established. This protocol allowed us to directly measure the effects of individual CCP transfusions on antibody levels in patients receiving CCP units with a wide range of antibody levels [18-20]. While usage of CCP has declined, 4 subjects in our study have now received CCP from vaccinated donors. This has allowed us to compare antibody responses with CCP from vaccinated and unvaccinated donors, providing data that could be used to design future studies related to the use of CCP for the treatment of COVID-19.

## Methods

### CCP Recipient protocol

Some COVID-19 patients were transfused under the expanded access protocol widely used in the United States that allowed for the transfusion of 1-2 units of CCP to hospitalized patients with severe or life threatening disease (NCT04338360). Other patients were treated under the emergency use authorization provided by the Food and Drug Administration (FDA). Blood bank staff informed study staff when CCP was issued to patients. Study personnel tracked the transfusion of the CCP, noting the starting and ending times for the CCP transfusion(s), and retrieved residual clinical samples (mint green top lithium heparin plasma separator tubes, PST) from the clinical core laboratory prior to discard. Plasma samples were aliquoted and frozen at -80C without any additional testing by research staff. No testing was performed until patient consent was obtained. For the 25 subjects included in this study, the pretransfusion samples were drawn from 5.3 to 15.6 hours (mean 9.7 / median 9.6) before transfusion. The samples obtained after the CCP transfusion were drawn from 1.6 to 16.8 hours (mean 10.5 / median 11.2) after the end of CCP transfusion. Patients were recruited under an IRB approved protocol (#202004503) that allowed study staff to explain the study by phone and/or email if necessary and allowed consent forms be sent electronically and by US mail if necessary.

### Patient Characteristics

Patient’s admission and discharge date, age, sex, weight, and plasma unit volume were collected from electronic health records. The twenty five subjects in this study had an average age of 55.9 (median= 59) and ranged in age from 27 to 87. Nine of the 25 subjects were females (36%). Twenty-two of the patients received CCP within three days of hospitalization and three received the plasma after 2 days. Twelve subjects received 1 unit (including 4 who each received one unit of CCP from vaccinated donors) and 13 were transfused with 2 units. The mean number of symptomatic days prior to CCP transfusion was 10.7 (median=10.5) with a range of 2 to 23 days to treatment. All the patients in the study were discharged with the average days to discharge from CCP transfusion of 8.1 days (median=4) and range from 1 to 41 days. Patient charesteristics and a summary of the antibody concentration of CCP they received is found in **Table 1**.

**Table 1.** CCP recipient information.

| Pt ID | Sex | Age | Weight (Kg) | CCP Spike IgG AU/ml | Days from Symptoms to CCP | Days to Discharge from CCP | Hosp Days to CCP |
| --- | --- | --- | --- | --- | --- | --- | --- |
| 1 | M | 60s | 105 | 104 | 2 | 1 | 1 |
| 2 | F | 30s | 166 | 71 | 4 | 7 | 2 |
| 3 | M | 80s | 62 | 30.5 | 15 | 19 | 1 |
| 4 | M | 50s | 104 | 74.3 | 16 | 1 | 0 |
| 5 | F | 20s | 66 | 105 | 13 | 8 | 5 |
| 6 | F | 60s | 51 | 47.3 | 8 | 2 | 1 |
| 7 | M | 40s | 78 | 27 | 14 | 1 | 2 |
| 8 | F | 60s | 90 | 30.5 | 10 | 9 | 0 |
| 9 | M | 60s | 100 | 124/308 | 8 | 41 | 2 |
| 10 | M | 60s | 71 | 124;159 | 2 | 3 | 2 |
| 11 | M | 60s | 92 | 114/288 | 2 | 8 | 1 |
| 12 | F | 60s | 77 | 52/136 | 9 | 15 | 1 |
| 13 | M | 50s | 93 | 52/136 | 5 | 8 | 1 |
| 14 | M | 40s | 69 | 60/71 | 12 | 7 | 1 |
| 15 | M | 50s | 104 | 21/52 | 10 | 1 | 0 |
| 16 | F | 50s | 89 | 132/375 | 12 | 1 | 2 |
| 17 | F | 40s | 82 | 16/131 | 22 | 2 | 0 |
| 18 | M | 60s | 79 | 110/163 | 13 | 4 | 2 |
| 19 | M | 70s | 113 | 159/196 | 1 | 2 | 1 |
| 20 | M | 50s | 116 | 37/38 | 4 | 6 | 2 |
| 21 | F | 60s | 142 | 31/47 | 23 | 39 | 19 |
| 22 | M | 40s | 159 | 4137 | 10 | 12 | 5 |
| 23 | F | 60s | 53 | 6783 | NA | 1 | 2 |
| 24 | M | 50s | 81 | 6783 | 7 | 1 | 1 |
| 25 | M | 20s | 80 | 3066 | NA | 4 | 1 |

### CCP donor screening and testing

CCP donors were screened following FDA guidance instructions under an IRB approved protocol (#202003554). The consent signed by all donors allowed the use of blood samples for research purposes. Donors were identified and screened following FDA guidelines at the time they enrolled. Donors either had PCR confirmed COVID-19 or had signs or symptoms of COVID-19 and were screened using serological testing. Serum from all subjects was stored at -80C for SARS-CoV-2 antibody testing as it became available. Samples for this study have been tested using DiaSorin Sars-CoV-2 IgG assay on the Liaison XL instrument (Stillwater MN) which has a positive cutoff of 15 arbitrary units per ml (AU/ml). DiaSorin’s high sensitivity (>95%) indicates that it is a promising assay for confirming prior SARS-CoV 2 infection [21]. A value of 30 AU/ml or higher was initially used to be eligible for CCP donation. As supply of CCP exceeded demand, a value of 100 AU/ml was used to allow donation of CCP. As donors started to get vaccinated, we concentrated our collection efforts on those donors who had been vaccinated. Antibody results from these donors have previously been reported and the average value for antibody positive unvaccinated donors was 82 AU/ml while the average antibody level from vaccinated donors was 4166 AU/ml [19].

### CCP dose and predicting SARS-CoV-2 antibody levels in recipients

For this study, the antibody dose provided to each patient was calculated using the donor/unit antibody level measured with the DiaSorin IgG assay (AU/ml) and multiplying it by the volume (L) of each unit (reported as kilo arbitrary units = kAU). The average antibody concentration in the 34 units from unvaccinated donors that were transfused as part of this study was 107 AU/ml and ranged from 21 to 375 AU/ml. The average dose of these units was 25.9 kAU and ranged from 4.5 to 97.5 kAU. The average antibody concentration in the 4 units from vaccinated donors that were transfused as part of this study was 5192 AU/ml and ranged from 3066 to 6783 AU/ml. The average dose of these 4 units was 1169 kAU and ranged from 782 to 1458 kAU. When two units of CCP were transfused, the dose was simply the sum of the dose for those two units. The predicted antibody level following transfusion was calculated as follows. The patient’s blood volume (L) was calculated from the patient’s sex, height, and weight following the calculations routinely used for apheresis [22]. The plasma volume (L) was then calculated by multiplying the blood volume x (100% Hematocrit). The predicted antibody level following transfusion was calculated by dividing the total CCP dose (kAU) by the patient’s plasma volume (L) and adding this to the patient’s SARS-CoV-2 antibody level (AU/ml) before transfusion.

## Results

### SARS-CoV-2 antibody testing in patients before and after transfusion

Twenty-five CCP recipients have been tested for SARS-CoV-2 antibodies before (pre) and after (post) the CCP transfusion, and results for those receiving just one unit of CCP are shown in Figure 1A. Of these subjects, 8 received CCP from unvaccinated donors while 4 received CCP from vaccinated donors. Of those receiving unvaccinated CCP, 6 were seronegative (<15 AU.ml) prior to the transfusion and just 2 of these were seropositive (≥15 AU/ml) after CCP transfusion. Of those receiving CCP from vaccinated donors, 3 were seronegative prior to the transfusion and all 3 were seropositive after transfusion. In those receiving CCP from vaccinated donors, the average post transfusion antibody level was 210 AU/ml, a value higher even than the average value seen in CCP donors who had recovered from COVID-19 [16]. The results for those receiving 2 units of CCP are shown in Figure 1B. Nine of the 12 patients in this group had negative antibody levels before transfusion and 7 were antibody-positive after transfusion. Of the 7 antibody negative patients who received 2 units of CCP, the highest post transfusion antibody level detected was 61 AU/ml. All but one of the patients receiving 2 units of CCP showed an increase in antibody levels and the one exception was a patient whose antibody level prior to transfusion was 131 AU/ml. This value was higher than both of the CCP units they received so it is not surprising that their antibody level was reduced slightly by the transfusions. These results demonstrate that 1 unit of CCP from vaccinated donors with very high antibody levels provides a much larger antibody increment than even 2 units of CCP from unvaccinated donors.

**Figure 1:**
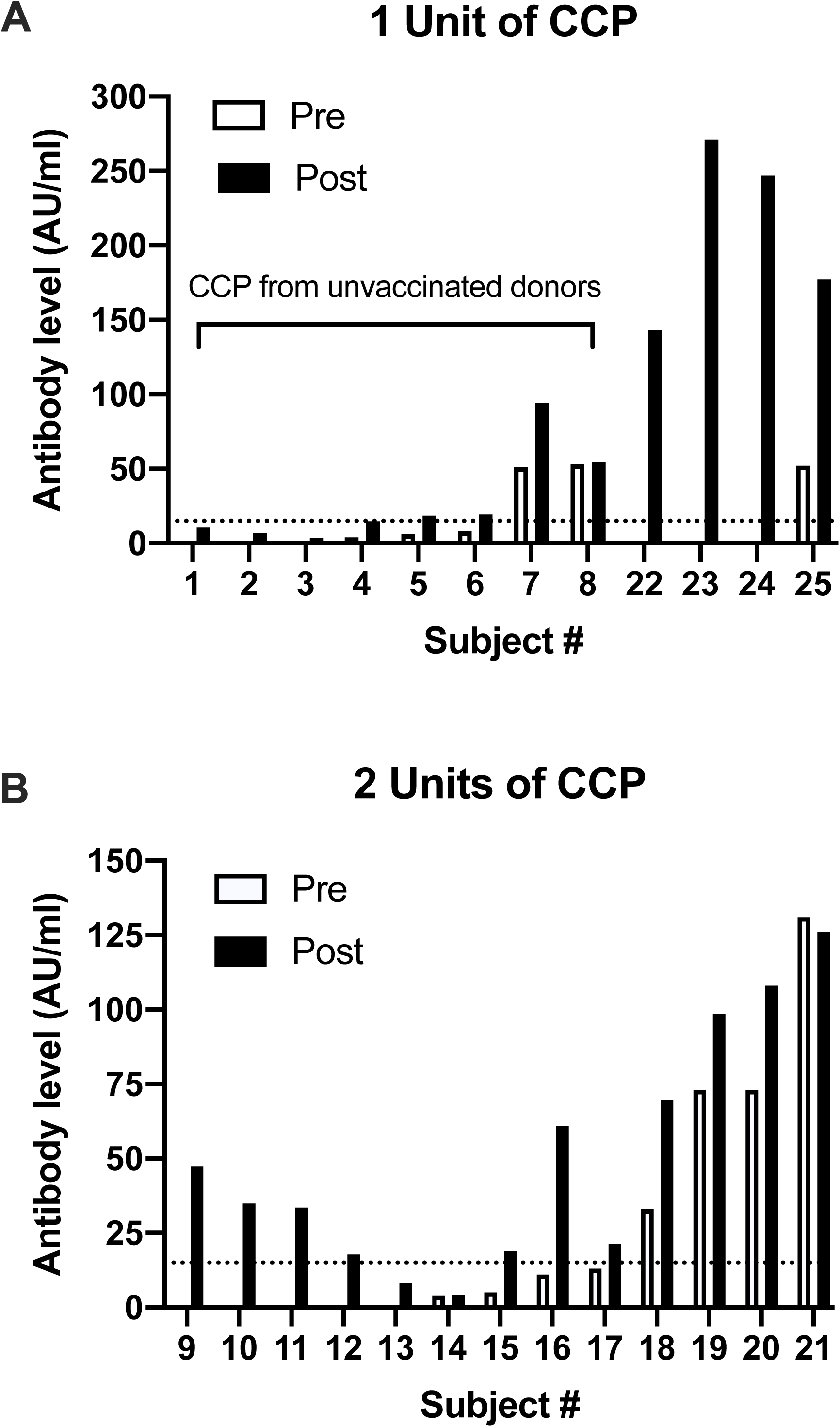
SARS-CoV-2 antibody levels in CCP recipient before (pre) and after (post) CCP transfusion. All samples were tested using the Diasorin IgG assay which has a positive cutoff of 15 AU/ml (dashed line). A) subjects who received 1 unit of plasma. Subjects 1-8 received CCP from unvaccinated donors while 22-25 received CCP from vaccinated donors. B) Subjects who received 2 units of CCP from unvaccinated donors.

### CCP “dose” and changes in antibody levels in CCP recipients

We hypothesized that the amount of antibody in the CCP would correlate strongly with changes in antibody levels in CCP recipients. The antibody dose was calculated by multiplying the donor antibody level (AU/ml) by the unit volume (L). If two units were given, then these were simply added together. Figure 2 shows that the CCP dose strongly correlates with the difference in the patient post-CCP antibody level minus the pre-level (R^2^ = 0.95). Of note, 7 of 7 seronegative patients receiving a dose greater than 50 kAU seroconverted after transfusion. In contrast, only 5 of 11 seronegative patients who received a CCP dose of less than 50 kAU seroconverted after transfusion.

**Figure 2:**
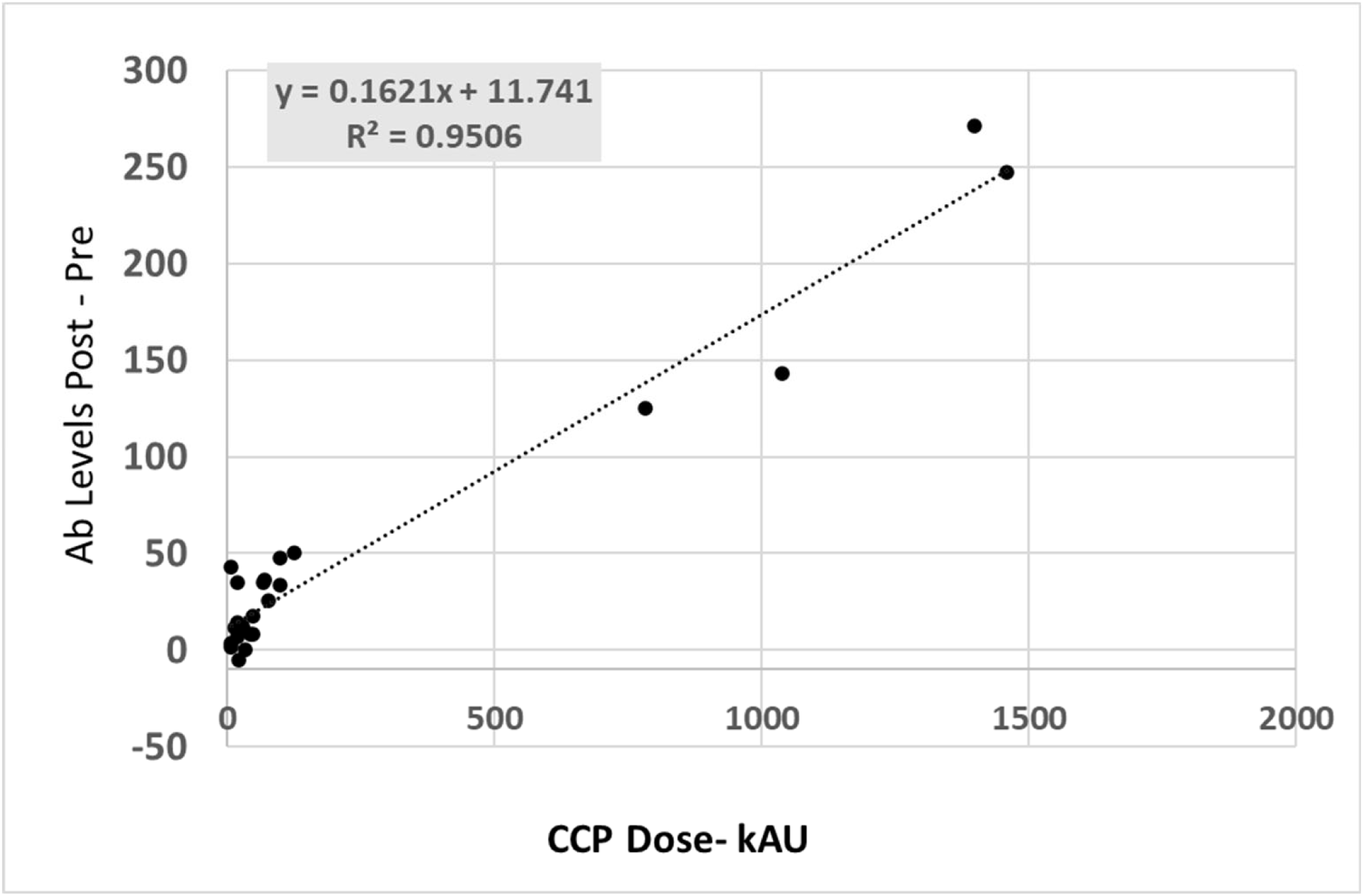
Relationship between changes in SARS-CoV-2 and CCP dose. Changes (post minus pre) in SARS-CoV-2 antibody levels (AU/ml) in CCP recipients is plotted against the dose of CCP which was calculated as described in materials and methods and reported as kilo arbitrary units (kAU).

### Predicting patient antibody level using CCP dose and patient plasma volume

We hypothesized that factoring in the patient’s plasma volume, patient antibody level prior to transfusion, and the CCP dose may be used to predict the patient’s antibody level after transfusion. For these calculations, the CCP dose (kAU) was divided by the patient’s plasma volume (L) and this was then added to the patient’s pre-transfusion antibody level (AU/ml). Figure 3 shows the predicted post-CCP vs actual post-CCP antibody levels for the patients in this study. This simple formula was surprisingly accurate in predicting these patient’s antibody levels (R^2^ = 0.92).

**Figure 3:**
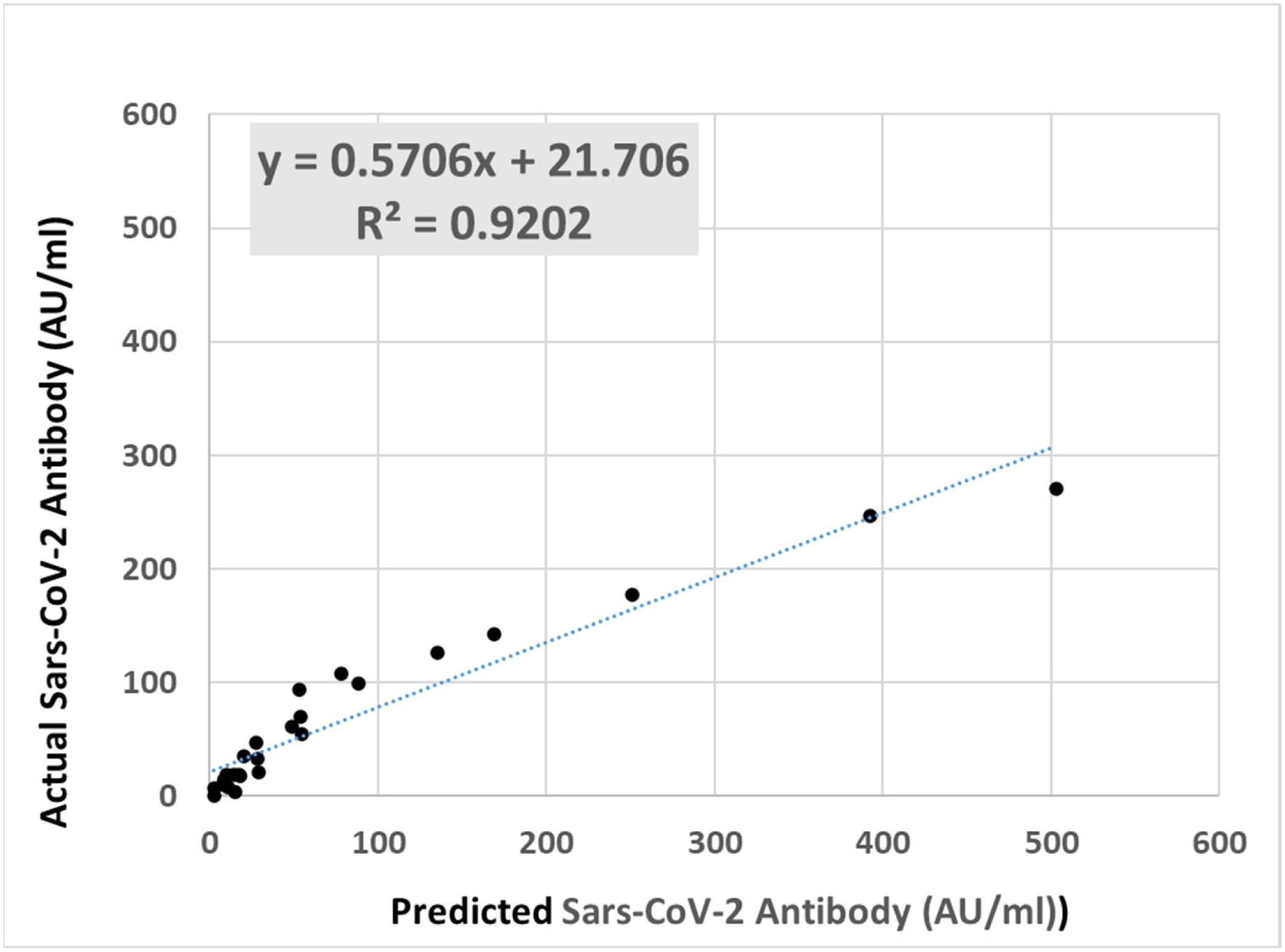
Predicting SARS-CoV-2 antibody levels in CCP recipients. Recipient plasma before and after transfusion were tested using the DiaSorin IgG assay. The predicted post transfusion antibody level (AU/ml) was calculated as described in the materials and methods and is plotted against the actual or measured antibody level in each subject.

## Discussion

As a CCP program was established at our donor center, we suggested that CCP facilities use existing SARS-CoV-2 serological assays to screen CCP donors and collect samples from recipients for future research use [20]. Using this approach we found that the CCP dose (kAU) is a strong predictor of changes in antibody level in recipients of CCP. One primary determinant of the dose is the number of units received by the patient. Patients receiving just one unit of CCP from unvaccinated donors showed modest changes in antibody levels, whereas patients receiving 2 units of CCP from unvaccinated donors were much more likely to seroconvert. Focusing on recipients who were seronegative before transfusion, in those receiving a CCP dose of 50 kAU or higher, all 7 of these patients seroconverted, providing some insight into a threshold antibody titer that could be targeted when selecting units for transfusion. During the course of this study, the antibody level required for donation of CCP was increased from 30 to 100 AU/ml. With a value of 100 AU/ml and a minimum volume of 0.25 liters, the minimum dose of one unit of CCP would be 25 kAU (.25 × 100). Thus this data would indicate that using 2 units of CCP from donors with antibody levels > 100 AU/ml would be very likely to seroconvert recipients. While patient specific results will certainly vary, this approach would provide a simple method of selecting CCP units with the highest probability of seroconversion following transfusion.

The FDA modified the CCP donor criteria to allow for collection of CCP from vaccinated donors on January 15, 2021. At this time we refocused our CCP collection efforts exclusively on vaccinated donors as we found these donors had exponentially higher antibody levels [16]. While nearly 20 vaccinated CCP donors have been tested to date, CCP was only collected from 6 due to donor restrictions outlined under the FDA guidance. Unfortunately, the donors who had the highest antibody responses (some in excess of 10,000 AU/ml) were more than 6 months out from recovery and therefore ineligible to donate CCP [16]. The most striking findings in this report are the dramatic changes in antibody levels in the patients who received CCP from vaccinated donors. All had antibody levels following transfusion in excess of 100 AU/ml which is higher than the average levels seen in donors who had recovered from COVID-19 and far higher than the levels seen in the patients receiving CCP from unvaccinated donors. Whether these improved antibody responses with CCP from vaccinated donors result in improved outcomes is not known. This determination would require a well-controlled clinical trial focused on CCP from vaccinated donors. To this point, the data from the expanded access protocol which showed that patients who received “high-titer” CCP had improved survival would certainly support the hypothesis that vaccinated CCP could be superior to CCP from unvaccinated donors [15]. Of note, none of the CCP infused under the expanded access protocol could have come from vaccinated donors as CCP from vaccinated donors was not allowed until January of 2021, well after these patients had been treated.

To our knowledge, this is the first study that systematically examined antibody responses in patients receiving CCP and compared those results to the antibody units in the donors. While the optimal antibody level in COVID-19 patients is not known, a reasonable initial “transfusion goal” would be to seroconvert these patients following transfusion [18]. Our results show that a single unit of CCP from an unvaccinated donor commonly is not sufficient to achieve this result as 4 of 6 of these recipients remained seronegative after transfusion. Using 2 units of unvaccinted CCP or one unit of vaccinated CCP was much more successful in seroconverting recipients with 7 of 9 recipients seroconverting following 2 units of unvaccinated CCP and 3 of 3 recipients of vaccinated CCP seroconverting. Another possible “goal” for antibody levels following CCP treatment might be to achieve antibody levels commonly observed in patients recovered from COVID-19. In our studies, we found that CCP donors following recovery had an average spike specific antibody level of 82 AU/ml [16]. No seronegative patients receiving unvaccinated CCP had antibody levels this high. In contrast, the three seronegative patients who received CCP from vaccinated donors all had antibody levels over 100 AU/ml. Of note, one of these subjects had an estimated blood volume of over 8 L so these results could apply to nearly all patients independent of their size. These results demonstrate that a study using CCP from vaccinated donors could consistently achieve antibody levels commonly seen after recovery from COVID-19.

## Data Availability

Deidentified data from this study will be available upon request.

## Acknowledgments

JL assisted in the design of the study, consented subjects and assisted in the collection of samples. SD and AE assisten in the collection of the plasma samples and facilitated the testing of these samples. JK assisted in the consenting of donors and cooridination in the collection of donor samples. AM, KR and JBJ assisted in the experimental design and the writing of the manuscript. CMK was the principle investigator for this study, participated in the experimental design, sample collection and primarily wrote the manuscript.

We would like to thank Dena Voss who assisted in the collection and analysis of residual samples from CCP recipients, staff involved in donor consenting and screening including Barb Swanson, Meredith Parsons, and Samantha Kouba; and staff at the state hygienic lab who assisted in testing CCP donors including Michelle Sexton, Haley Peden, and Michael Pentella.

